# Effect of seizures on developmental trajectories in children with autism

**DOI:** 10.1101/2022.03.30.22273179

**Authors:** Phillip Forman, Edward Khokhlovich, Andrey Vyshedskiy

**Author notes:** Corresponding author: Andrey Vyshedskiy, Ph.D., Boston University, Boston, USA.

## Abstract

The effect of seizures in 2 to 5-year-old children with ASD was investigated in the largest and the longest observational study to-date. Parents assessed the development of 8461 children quarterly for three years on five orthogonal subscales: combinatorial receptive language, expressive language, sociability, sensory awareness, and health. Seizures were reported in 11% of children. Children with no seizures developed faster compared to matched children with seizures in all subscales. On an annualized basis, participants with no seizures improved their receptive language 1.5-times faster than those with seizures; expressive language: 1.3-times faster; sociability: 2.3-times faster; sensory awareness: 6.2-times faster; and health: 20.0-times faster. This study confirms a high prevalence of seizures in ASD children and points to the need for more systematic approach to the development of treatment strategies.

## Introduction

Seizures are known to negatively affect children cognitive and language development (Elger et al., 2004; Holmes, 2015). Schoenfeld et al. found that children with seizures performed 14.9% worse than sibling control group on the reading subscale (p<0.001), 16.3% worse on the spelling subscale (*p*<0.001), and 12.8% worse on the math subscale of the Wide Range Achievement Test (*p*<0.01) (Schoenfeld et al., 1999). They also had 10.6% lower performance on K-BIT Expressive Vocabulary test (*p*<0.001), 27.9% lower on the Controlled Oral Word Association Test (*p*<0.001), 21.3% lower on the Category Fluency test (*p*<0.001), and 8.0% lower on the Children’s Token Test (*p*<0.001) (Schoenfeld et al., 1999). Hermann et al. found that children with early onset of seizures scored 11.6% lower on WAIS-III Verbal IQ scores (*p*<0.0001), 11.3% lower on the Boston Naming Test (*p*<0.0001), and 21.2% lower on the Controlled Oral Word Fluency test (*p*=0.002) as compared to a healthy age-matched controls (Hermann et al., 2002). Esmael et al. found that children with selective language impairments had a 26.2% prevalence of abnormal EEGs compared to 6.2% in the age- and sex-matched control group (*p*=0.006) (Esmael et al., 2021). There was a strong negative correlation between language intelligence quotient and abnormal EEGs (r=-0.91, p<0.01).

Most studies, however, suffer from low numbers of participants and lack of longitudinal data. In 2015 we published a language training app for children (Dunn, Elgart, Lokshina, Faisman, Khokhlovich, et al., 2017b, 2017a; Dunn, Elgart, Lokshina, Faisman, Waslick, et al., 2017; Vyshedskiy et al., 2020; Vyshedskiy & Dunn, 2015), which invites parents to complete their child’s evaluation and seizure status every three months. As a result, we accumulated over a hundred thousand longitudinal evaluations. This gives us an opportunity to study children’s developmental trajectories. In this manuscript, we describe the investigation of the effect of seizures as reported by parents. To assess the effect of seizures, we compared participants with no seizures to participants with mild, moderate, and serious seizures.

Expressive language, sociability, sensory awareness, and health were assessed by Autism Treatment Evaluation Checklist (ATEC) (Rimland & Edelson, 1999), a measure validated for longitudinal tracking of symptoms and assessing changes in ASD severity (Charman et al., 2004; Klaveness et al., 2013; Magiati et al., 2011; Mahapatra, Khokhlovich, et al., 2018). The expressive language subscale of ATEC consists of 14 items; the sociability subscale contains 20 items; the sensory awareness subscale has 18 items; and the health subscale contains 25 items.

Combinatorial receptive language was assessed with Mental Synthesis Evaluation Checklist (MSEC) (Braverman et al., 2018), a 20-item questionnaire that focuses on combinatorial language skills. MSEC items include questions like: Understands simple stories that are read aloud; Understands elaborate fairy tales that are read aloud; Understands some simple modifiers; Understands several modifiers in a sentence; Understands size; Understands spatial prepositions; Understands the change in meaning when the order of words is changed; Understands explanations about people, objects or situations beyond the immediate surroundings; and so on. MSEC has been shown to have good internal reliability, adequate test– retest reliability, good construct validity, and good known group validity (Braverman et al., 2018).

The framework for the evaluation of score changes over time was developed in Mahapatra *et al*. (Mahapatra, Khokhlovich, et al., 2018) and further validated in Vyshedskiy *et al*. (Vyshedskiy et al., 2020) and Fridberg *et al*. (Fridberg et al., 2021). This robust technique uses the Linear Mixed Effect Model with Repeated Measures to model complex relationships between time of evaluation, group assignment (no Seizures Group or Seizures Group), and evaluation subscales. The model then computes averages for each group and for each subscale at each time point.

## Methods

### Participants

Participants were users of a language therapy app that was made available gratis at all major app stores in September 2015. Once the app was downloaded, caregivers were asked to register and to provide demographic details, including the child’s diagnosis and age. Caregivers consented to anonymized data analysis and completed the Autism Treatment Evaluation Checklist (ATEC) (Rimland & Edelson, 1999), an evaluation of the receptive language using the Mental Synthesis Evaluation Checklist (MSEC) (Braverman et al., 2018), as well as the Screen Time assessment and the Diet and Supplements assessment. The first evaluation was administered approximately one month after the download. The subsequent evaluations were administered at approximately three-month intervals. To enforce regular evaluations, the app became unusable at the end of each three-month interval and parents were required to complete an evaluation to regain its functionality.

### Inclusion criteria

Inclusion criteria were identical to our previous studies of this population (Fridberg et al., 2021; Mahapatra, Khokhlovich, et al., 2018; Vyshedskiy et al., 2020). Specifically, we selected participants based on the following criteria:

1. Consistency: Participants must have filled out at least three ATEC evaluations and the interval between the first and the last evaluation was six months or longer.
2. Diagnosis: ASD. Children without ASD diagnosis were excluded from the study. Other diagnostic options included: Mild Language Delay, Pervasive Developmental Disorder, Attention Deficit Disorder, Social Communication Disorder, Specific Language Impairment, Apraxia, Sensory Processing Disorder, Down Syndrome, Lost Diagnosis of Autism or PDD, Other Genetic Disorder, normally developed child. The parent-reported ASD diagnosis was supported by ATEC scores. Average initial ATEC total score was 68.5 ± 25.1, which corresponds to moderate-to-severe ASD as delineated earlier (Mahapatra, Vyshedsky, et al., 2018) and Table 1.

**Table 1.**
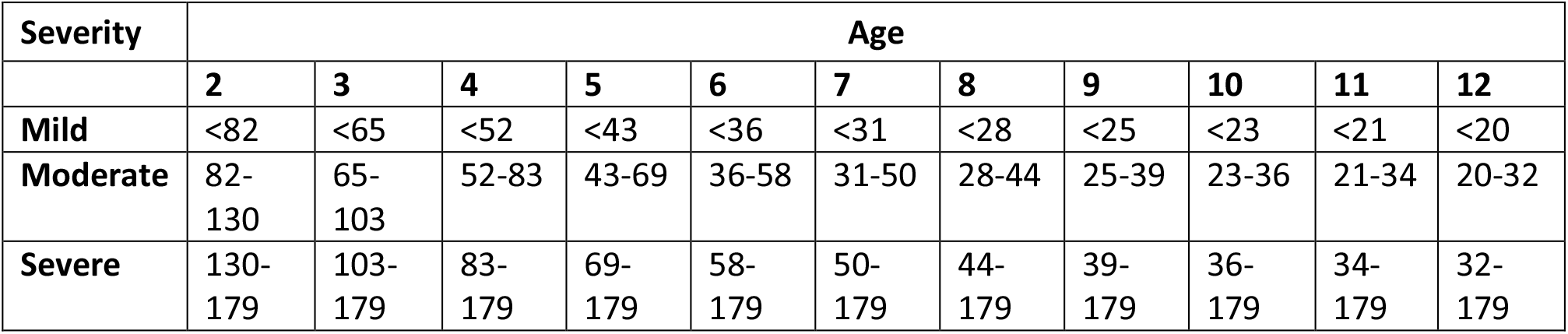
Approximate relationship between ATEC total score, age, and ASD severity as described in Mahapatra *et al*. (Mahapatra, Khokhlovich, et al., 2018). At any age, a greater ATEC score indicates greater ASD severity.

| Severity | Age |  |  |  |  |  |  |  |  |  |  |
| --- | --- | --- | --- | --- | --- | --- | --- | --- | --- | --- | --- |
|  | 2 | 3 | 4 | 5 | 6 | 7 | 8 | 9 | 10 | 11 | 12 |
| **Mild** | <82 | <65 | <52 | <43 | <36 | <31 | <28 | <25 | <23 | <21 | <20 |
| **Moderate** | 82-130 | 65-103 | 52-83 | 43-69 | 36-58 | 31-50 | 28-44 | 25-39 | 23-36 | 21-34 | 20-32 |
| **Severe** | 130-179 | 103-179 | 83-179 | 69-179 | 58-179 | 50-179 | 44-179 | 39-179 | 36-179 | 34-179 | 32-179 |

### Exclusion criteria

1. Maximum age: Participants older than five years of age at the time of their first evaluation were excluded from this study.
2. Minimum age: Participants who completed their first evaluation before the age of two years were excluded from this study.

After excluding participants that did not meet these criteria, there were 8461 total participants.

### Seizures group assessment

Participants were required to assess seizure problems while responding to the enquiry. The possible answers were: “not a problem,” “minor problem,” “moderate problem,” “serious problem.” To assess the effect of seizures, we compared participants with no reported seizures (N=7503; the no seizures group, abbreviated noSG) to participants with reported minor, moderate and serious seizure problem (N=958; the seizures group, abbreviated SG).

### Evaluations

A caregiver-completed Autism Treatment Evaluation Checklist (ATEC) (Rimland & Edelson, 1999) and Mental Synthesis Evaluation Checklist (MSEC) (Braverman et al., 2018) were used to track child development. The ATEC questionnaire is comprised of four subscales: 1) Speech/Language/Communication, 2) Sociability, 3) Sensory/Sensory awareness, and 4) Physical/Health/Behavior. The first subscale, Speech/Language/Communication, contains 14 items and its score ranges from 0 to 28 points. The Sociability subscale contains 20 items within a score range of 0 to 40 points. The third subscale, referred here as the Sensory awareness subscale, has 18 items and score range from 0 to 36 points. The fourth subscale, referred here as the Health subscale, contains 25 items and score range from 0 to 75 points. The scores from each subscale are combined in order to calculate a Total Score, which ranges from 0 to 179 points. A lower score indicates lower severity of ASD symptoms and a higher score indicates more severe symptoms of ASD. ATEC is not a diagnostic checklist and it was designed to evaluate the effectiveness of treatment ^16^. Therefore, ASD severity can only have an approximate relationship with the total ATEC score and age. Table 1 lists approximate ATEC total score as related to ASD severity and age as described in Mahapatra *et al*. (Mahapatra, Khokhlovich, et al., 2018).

ATEC was selected as a tool since it is one of the few measures validated to evaluate treatment effectiveness. In contrast, another popular ASD assessment tool, Autism Diagnostic Observation Schedule or ADOS, (Lord et al., 2000) has only been validated as a diagnostic tool. Various studies confirmed the validity and reliability of ATEC (Al Backer, 2016; Geier et al., 2013; Jarusiewicz, 2002) and several trials confirmed ATEC’s ability to longitudinally measure changes in participant performance (Charman et al., 2004; Klaveness et al., 2013; Magiati et al., 2011; Mahapatra, Khokhlovich, et al., 2018). Moreover, ATEC has been used as a primary outcome measure for a randomized controlled trial of iPad-based intervention for ASD, named “Therapy Outcomes By You” or TOBY, and it was noted that ATEC’s possesses an “internal consistency and adequate predictive validity” (Whitehouse et al., 2017). These studies support the effectiveness of ATEC as a tool for longitudinal tracking of symptoms and assessing changes in ASD severity.

### Expressive language assessment

The ATEC Speech/Language/Communication subscale includes the following questions: 1) Knows own name, 2) Responds to ‘No’ or ‘Stop’, 3) Can follow some commands, 4) Can use one word at a time (No!, Eat, Water, etc.), 5) Can use 2 words at a time (Don’t want, Go home), 6) Can use 3 words at a time (Want more milk), 7) Knows 10 or more words, 8) Can use sentences with 4 or more words, 9) Explains what he/she wants, 10) Asks meaningful questions, 11) Speech tends to be meaningful/relevant, 12) Often uses several successive sentences, 13) Carries on fairly good conversation, and 14) Has normal ability to communicate for his/her age. With the exception of the first three items, all items in the ATEC subscale 1 primarily measure expressive language. Accordingly, the ATEC subscale 1 is herein referred to as the Expressive Language subscale to distinguish it from the Receptive Language subscale tested by the MSEC evaluation.

### Receptive language assessment

The MSEC evaluation was designed to be complementary to ATEC in measuring complex receptive language. Out of 20 MSEC items, those that directly assess receptive language are the following: 1) Understands simple stories that are read aloud; 2) Understands elaborate fairy tales that are read aloud (i.e., stories describing FANTASY creatures); 3) Understands some simple modifiers (i.e., green apple vs. red apple or big apple vs. small apple); 4) Understands several modifiers in a sentence (i.e., small green apple); 5) Understands size (can select the largest/smallest object out of a collection of objects); 6) Understands possessive pronouns (i.e. your apple vs. her apple); 7) Understands spatial prepositions (i.e., put the apple ON TOP of the box vs. INSIDE the box vs. BEHIND the box); 8) Understands verb tenses (i.e., I will eat an apple vs. I ate an apple); 9) Understands the change in meaning when the order of words is changed (i.e., understands the difference between ‘a cat ate a mouse’ vs. ‘a mouse ate a cat’); 10) Understands explanations about people, objects or situations beyond the immediate surroundings (e.g., “Mom is walking the dog,” “The snow has turned to water”). MSEC consists of 20 questions within a score range of 0 to 40 points; similarly to ATEC, a lower MSEC score indicates a better developed receptive language.

The psychometric quality of MSEC was tested with 3,715 parents of ASD children (Braverman et al., 2018). Internal reliability of MSEC was good (Cronbach’s alpha > 0.9). MSEC exhibited adequate test–retest reliability, good construct validity, and good known group validity as reflected by the difference in MSEC scores for children of different ASD severity levels.

To simplify interpretation of figure labels, the subscale 1 of the ATEC evaluation is herein referred to as the Expressive Language subscale and the MSEC scale is referred to as the Receptive Language subscale.

### Statistical analysis

The framework for the evaluation of score changes over time has been earlier explained in detail in Mahapatra *et al*. (Mahapatra, Khokhlovich, et al., 2018) and Vyshedskiy *et al*. (Vyshedskiy et al., 2020). In short, the concept of a “Visit” was developed by dividing the three-year-long observation interval into 3-month periods. All evaluations were mapped into 3-month-long bins with the first evaluation placed in the first bin. When more than one evaluation was completed within a bin, their results were averaged to calculate a single number representing this 3-month interval. Thus, we had 12 quarterly evaluations for both noSG and SGs.

It was then hypothesized that there was a two-way interaction between pretend-play-group and Visit. Statistically, this hypothesis was modeled by applying the Linear Mixed Effect Model with Repeated Measures (MMRM), where a two-way interaction term was introduced to test the hypothesis. The model (Endpoint ∼ Baseline + Gender + Severity + Seizures-Group * Visit) was fit using the R Bioconductor library of statistical packages, specifically the “nlme” package. The subscale score at baseline, as well as gender and severity were used as covariates. Conceptually, the model fits a plane into n-dimensional space. This plane considers a complex variability structure across multiple visits, including baseline differences. Once such a plane is fit, the model calculates Least Squares Means (LS Means) for each subscale and group at each visit. The model also calculates LS Mean differences between the groups at each visit.

To control for the health score, a second Linear Mixed Effect Model was used: (Endpoint ∼ Baseline + Gender + Severity + Health + Seizures-Group * Visit). These results are reported separately.

In preparation for statistical analysis, participants in the SG were matched to those in the noSG using propensity score (Schneider et al., 2007) based on age, gender, expressive language, receptive language, sociability, Sensory awareness, and health at the 1^st^ evaluation (baseline). The number of matched participants was 643 in each group, Table 2.

**Table 2.** Demographic data of participant pool.

|  | No seizures | Seizures Group |
| --- | --- | --- |
| **Number of participants** | 7503 | 958 |
| **Number of matched participants** | 955 | 955 |
| **Age at baseline** | 3.5±0.8 | 3.5±0.8 |
| **Male Gender** | 74% | 74% |

## Results

Seizures were reported by 11% of participants (the Seizures Group, abbreviated SG). These participants were matched to those with no seizures (no Seizures Group, abbreviated noSG) using propensity score (Schneider et al., 2007) based on age, gender, expressive language, receptive language, sociability, Sensory awareness, and health at the 1^st^ evaluation (baseline). The number of matched participants was 955 out of 958 in the SG and 955 out of 7503 in the noSG. We then analyzed trajectories of children development on five orthogonal subscales: Receptive Language, Expressive Language, Sociability, Sensory awareness, and Health.

On the Receptive Language subscale, the average improvement in the noSG over 36 months was 6.07 points (SE=0.96, p<0.0001) compared to 4.12 points (SE=1.01, p<0.0001) in the SG, Figure 1A, Table 3, Table S1. The difference in the noSG relative to the SG at Month 36 was not statistically significant: -0.43 points (SE=1.38, p=0.7537). The negative difference (marked in the Table 3 as “noSG – SG”) indicates that the SG had greater scores at Month 36 and, therefore, more severe symptoms. On an annualized basis, the participants from noSG improved their receptive language 1.5-times faster than those in SG (noSG =2.0 points/year; SG =1.4 points/year).

**Table 3.** Characteristics of no Seizures Group (noSG) and Seizures Group (SG). Data is presented as LS Means (SE; 95% CI). A lower score indicates a lower severity of ASD symptoms. The difference between no Seizures Group and Seizures Group (noSG –SG) is presented as: LS Mean (SE; P-value). The negative noSG – SG difference indicates that SG had higher score and therefore more severe symptoms.

| Subscale | Baseline |  |  | Month 36 |  |  | Month 36 – Baseline |  |
| --- | --- | --- | --- | --- | --- | --- | --- | --- |
|  | No Seizures Group | Seizures Group | noSG – SG | No Seizures Group | Seizures Group | noSG – SG | No Seizures Group | Seizures Group |
| **Receptive Language** | 28<br>(0.246;<br>27.5 -<br>28.5) | 26.5 (0.261;<br>26 - 27) | 1.52<br>(0.34;<br><0.0001) | 21.9<br>(0.949;<br>20.1 -<br>23.8) | 22.4<br>(1.007;<br>20.4 -<br>24.3) | -0.43<br>(1.38;<br>0.7537) | -6.07<br>(0.96;<br><0.0001) | -4.12 (1.01;<br><0.0001) |
| **Expressive Language** | 16.23<br>(0.165;<br>15.91 -<br>16.6) | 17.8 (0.188;<br>17.43 -<br>18.2) | 0.41<br>(0.23;<br>0.0725) | 9.02<br>(0.619;<br>7.8 - 10.2) | 11.8<br>(0.748;<br>10.3 -<br>13.2) | -2.61 (0.9;<br>0.0037) | -7.21<br>(0.62;<br><0.0001) | -4.2 (0.66;<br><0.0001) |
| **Sociability** | 17<br>(0.207;<br>16.62 -<br>17.4) | 16.5 (0.22;<br>16.08 -<br>16.9) | 0.52<br>(0.29;<br>0.0692) | 11.5<br>(0.827;<br>9.87 -<br>13.1) | 14.1<br>(0.877;<br>12.34 -<br>15.8) | -2.58 (1.2;<br>0.0320) | -5.54<br>(0.84;<br><0.0001) | -2.44 (0.88;<br>0.0058) |
| **Sensory awareness** | 17.1<br>(0.182;<br>16.8 -<br>17.5) | 16.3 (0.194;<br>15.9 - 16.7) | 0.84<br>(0.25;<br>0.0008) | 12.6<br>(0.722;<br>11.2 - 14) | 15.5<br>(0.766; 14<br>- 17) | -2.97<br>(1.05;<br>0.0047) | -4.54<br>(0.73;<br><0.0001) | -0.73 (0.77;<br>0.3419) |
| **Health** | 28.7<br>(0.352;<br>28 - 29.4) | 28.4 (0.375;<br>27.7 - 29.2) | 0.25<br>(0.49;<br>0.6033) | 19.7<br>(1.393; 17<br>- 22.4) | 28 (1.479;<br>25.1 -<br>30.9) | -8.29<br>(2.02;<br><0.0001) | -8.99<br>(1.41;<br><0.0001) | -0.45 (1.49;<br>0.7633) |

**Figure 1.**
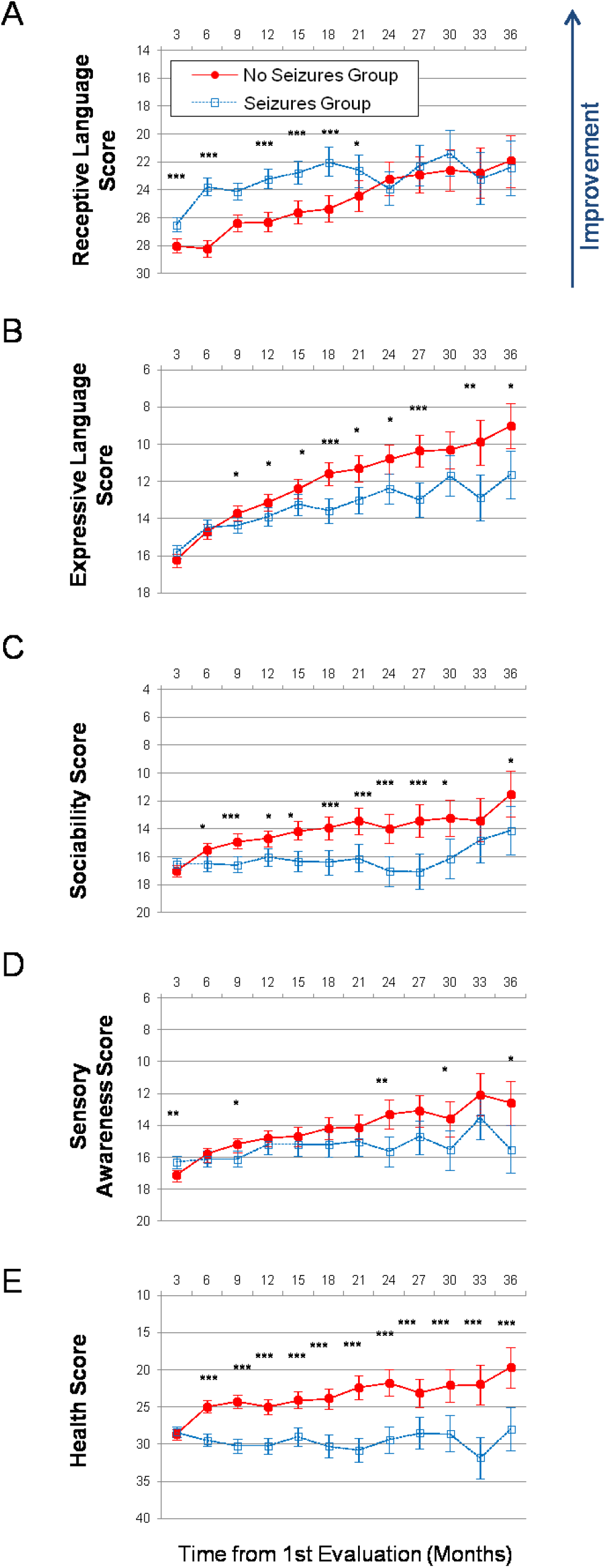
Longitudinal plots of subscale scores LS Means. Horizontal axis shows months from the 1^st^ evaluation (0 to 36 months). Error bars show the 95% confidence interval. To facilitate comparison between subscales, all vertical axes ranges have been normalized to show 40% of their corresponding subscale’s maximum available score. A lower score indicates symptom improvement. P-value is marked: ***<0.0001; **<0.001; *<0.05. (A) Receptive Language score. (B) Expressive Language score. (C) Sociability score. (D) Sensory Awareness score (E) Health score.

On the Expressive Language subscale, the participants in the noSG improved over the 36-month period by 7.21 points (SE=0.62, p<0.0001) compared to 4.2 points (SE=0.66, p<0.0001) improvement in the SG, Figure 1B, Table S2. The difference in the noSG relative to the SG at Month 36 was statistically significant: -2.61 points (SE=0.90, p=0.0037). On an annualized basis, the subjects in noSG improved their expressive language 1.3-times faster than those in SG (noSG =2.7 points/year; SG =2.0 points/year).

On the Sociability subscale, the subjects in the noSG improved over the 36-month period by 5.54 points (SE=0.84, p<0.0001) compared to 2.44 points (SE=0.88, p=0.0058) improvement in those in the SG, Figure 1C, Table S3. The difference in the noSG relative to the SG at Month 36 was statistically significant: -2.58 points (SE=1.2, p=0.0320). On an annualized basis, the participants in the noSG improved their sociability 2.3-times faster than those in the SG (noSG =1.9 points/year; SG =0.8 points/year).

On the Sensory awareness subscale, the subjects in the noSG improved over the 36-month period by 4.54 points (SE=0.73, p<0.0001) compared to 0.73 points (SE=0.77, p=0.3419) improvement in the SG, Figure 1D, Table S4. The difference in the noSG relative to the SG at Month 36 was statistically significant: -2.97 points (SE=1.05, p=0.0047). On an annualized basis, the participants in the noSG improved their sensory awareness 6.2-times faster than those in SG (noSG =1.5 points/year; SG =0.2 points/year).

On the Health subscale, the subjects in the noSG improved over the 36-month period by 8.99 points (SE=1.41, p<0.0001) compared to 0.45 points (SE=1.49, p=0.7633) in the SG, Figure 1E, Table S5. The difference in the noSG relative to the SG at Month 36 was statistically significant: -8.29 points (SE=2.02, p<0.0001). On an annualized basis, the participants in the noSG improved their health score 20.0-times faster than those in noSG (noSG =3.0 points/year; SG =0.2 points/year).

In order to investigate if the overall health was a driving force behind the group differences, we have re-run the model while controlling for the health score (in addition to each subscale score at baseline as well as gender and severity), Figure 2, Table 4. Controlling for the health score did not result in a significant change in the effect of seizures on any subscale. On the Receptive Language subscale, the average improvement in the noSG over 36 months was 7.31 points (SE=0.94, p<0.0001) compared to 3.91 points (SE=0.99, p<0.0001) in the SG, Figure 2A, Table 4, Table S6. The difference in the noSG relative to the SG at Month 36 was not statistically significant: -1.78 points (SE=1.35, p=0.1864). On an annualized basis, the participants from noSG improved their receptive language 1.9-times faster than those in SG (noSG =2.4 points/year; SG =1.3 points/year).

**Table 4.** Characteristics of no Seizures Group (noSG) and Seizures Group (SG) when controlling for the health score. Data is presented as LS Means (SE; 95% CI). A lower score indicates a lower severity of ASD symptoms. The difference between no Seizures Group and Seizures Group (noSG – SG) is presented as: LS Mean (SE; P-value). The negative noSG – SG difference indicates that SG had higher score and therefore more severe symptoms.

| Subscale | Baseline |  |  | Month 36 |  |  | Month 36 – Baseline |  |
| --- | --- | --- | --- | --- | --- | --- | --- | --- |
|  | No Seizures Group | Seizures Group | noSG – SG | No Seizures Group | Seizures Group | noSG – SG | No Seizures Group | Seizures Group |
| **Receptive Language** | 26.9<br>(0.286;<br>26.3 -<br>27.4) | 25.2 (0.298;<br>24.7 - 25.8) | 1.62<br>(0.33;<br><0.0001) | 19.5<br>(0.944;<br>17.7 -<br>21.4) | 21.3<br>(0.995;<br>19.4 -<br>23.3) | -1.78<br>(1.35;<br>0.1864) | -7.31<br>(0.94;<br><0.0001) | -3.91 (0.99;<br><0.0001) |
| **Expressive Language** | 16.24<br>(0.195;<br>15.86 -<br>16.6) | 15.89<br>(0.204;<br>15.49 -<br>16.3) | 0.35<br>(0.23;<br>0.1232) | 9.42<br>(0.631;<br>8.18 -<br>10.7) | 11.73<br>(0.665;<br>10.43 -<br>13) | -2.32 (0.9;<br>0.0103) | -6.83<br>(0.63;<br><0.0001) | -4.16 (0.66;<br><0.0001) |
| **Sociability** | 18.1<br>(0.237;<br>17.6 -<br>18.5) | 17.7 (0.246;<br>17.3 - 18.2) | 0.33<br>(0.28;<br>0.2359) | 14.4 (0.8;<br>12.8 -<br>15.9) | 15.6<br>(0.844;<br>13.9 -<br>17.2) | -1.2 (1.14;<br>0.294) | -3.71 (0.8;<br><0.0001) | -2.18 (0.84;<br>0.0097) |
| **Sensory awareness** | 16.9<br>(0.216;<br>16.5 -<br>17.4) | 16.1 (0.226;<br>15.7 - 16.6) | 0.83<br>(0.25;<br>0.001) | 12.5<br>(0.733;<br>11.1 - 14) | 15.7<br>(0.773;<br>14.2 -<br>17.2) | -3.13<br>(1.05;<br>0.0029) | -4.4 (0.73;<br><0.0001) | -0.44 (0.77;<br>0.5652) |

**Figure 2.**
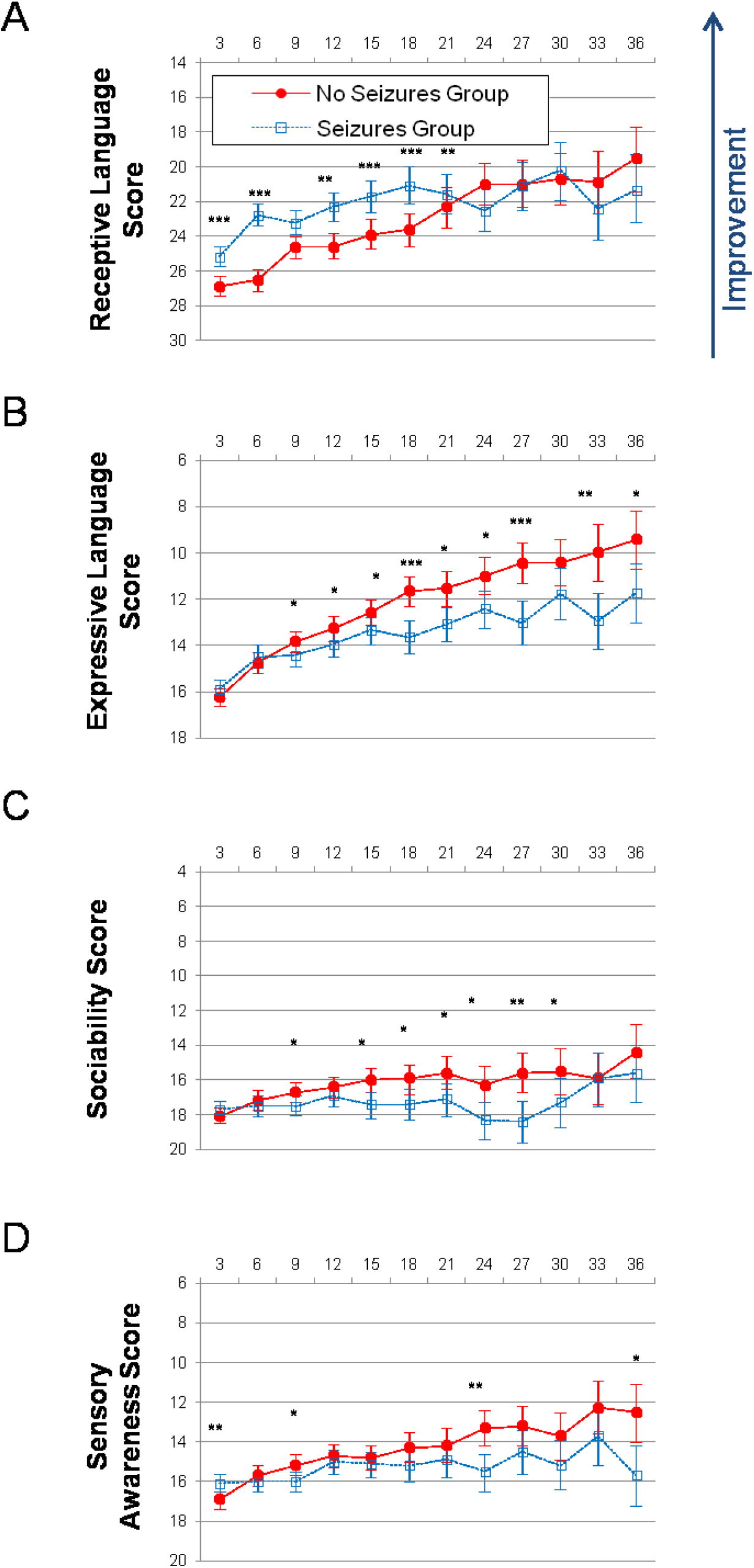
Longitudinal plots of subscale scores LS Means when controlling for the health score. Horizontal axis shows months from the 1^st^ evaluation (0 to 36 months). Error bars show the 95% confidence interval. To facilitate comparison between subscales, all vertical axes ranges have been normalized to show 40% of their corresponding subscale’s maximum available score. A lower score indicates symptom improvement. P-value is marked: ***<0.0001; **<0.001; *<0.05. (A) Receptive Language score. (B) Expressive Language score. (C) Sociability score. (D) Sensory awareness score.

On the Expressive Language subscale, the participants in the noSG improved over the 36-month period by 6.83 points (SE=0.63, p<0.0001) compared to 4.16 points (SE=0.66, p<0.0001) improvement in the SG, when controlling for the health score (in addition to the expressive language score at baseline as well as gender and severity), Figure 2B, Table S7. The difference in the noSG relative to the SG at Month 36 was statistically significant: -2.32 points (SE=0.90, p=0.0103). On an annualized basis, the subjects in noSG improved their expressive language 1.6-times faster than those in SG (noSG =2.3 points/year; SG =1.4 points/year).

On the Sociability subscale, the subjects in the noSG improved over the 36-month period by 3.71 points (SE=0.8, p<0.0001) compared to 2.18 points (SE=0.84, p=0.0097) improvement in those in the SG, when controlling for the health score (in addition to the expressive language score at baseline as well as gender and severity), Figure 2C, Table S8. The difference in the noSG relative to the SG at Month 36 was not statistically significant: -1.2 points (SE=1.14, p=0.294). On an annualized basis, the participants in the noSG improved their sociability 1.7-times faster than those in the SG (noSG =1.2 points/year; SG =0.7 points/year).

On the Sensory awareness subscale, the subjects in the noSG improved over the 36-month period by 4. 4 points (SE=0.73, p<0.0001) compared to 0.44 points (SE=0.77, p=0.5652) improvement in the SG, when controlling for the health score (in addition to the expressive language score at baseline as well as gender and severity), Figure 2D, Table S9. The difference in the noSG relative to the SG at Month 36 was: -3.13 points (SE=1.05, p=0.0029). On an annualized basis, the participants in the noSG improved their sensory awareness 10-times faster than those in SG (noSG =1.5 points/year; SG =0.15 points/year).

## Discussion

This is the largest and longest study to date demonstrating a strong association between seizures and children’s developmental trajectories. Over the period of three years, parents have quarterly assessed the development of 8461 two-to five-year-old children with ASD. Seizures were reported in 11% of individuals with ASD, in line with published prevalence of 2.4% to 26% (El Achkar & Spence, 2015; Viscidi et al., 2013). The ‘no Seizures Group’ (noSG) included 7503 children with no seizures and the ‘Seizures Group’ (SG) included 958 children with mild, moderate, and severe seizures. In order to compare the groups, participants in the SG group were matched to those in the noSG group using propensity score (Schneider et al., 2007) based on age, gender, expressive language, receptive language, sociability, sensory awareness, and health at the 1^st^ evaluation (baseline). The number of matched participants was 955 in each group.

Participants in the noSG improved to a greater extent than those in the SG in all subscales. On an annualized basis, the noSG participants improved their receptive language 1.5-times faster than SG; expressive language: 1.3-times faster; sociability: 2.3-times faster; sensory awareness: 6.2-times faster; and health score: 20.0-times faster. The difference reached statistical significance in the Combinatorial Receptive Language subscale on Months 3, 6, 12, 15, 18, and 21; in the Expressive Language subscale on Months 9 through 36 (with the exception of Month 30); in the Sociability subscale on Months 6 through 36 (with the exception of Month 30); in the Sensory awareness subscale on Months 3, 9, 24, 30, and 36; in the Health subscale on all time points except month 3. The results of this study support previous reports of a negative correlation between the presence of seizures and every aspect of a child’s development (Esmael et al., 2021; Hermann et al., 2002; Schoenfeld et al., 1999).

The greatest negative effect of seizures in our study was on the overall health of a child. Controlling for the health score (in addition to each subscale score at baseline as well as gender and severity) did not result in significant change of the effect of seizures on any subscale. On an annualized basis, the noSG participants improved their receptive language 1.9-times faster than SG; expressive language: 1.6-times faster; sociability: 1.7-times faster; sensory awareness: 10-times faster. The difference reached statistical significance in the Combinatorial Receptive Language subscale on Months 3, 6, 12, 15, 18, and 21; in the Expressive Language subscale on Months 9 through 36 (with the exception of Month 30); in the Sociability subscale on Month 9 and Months 12 through 30; in the Sensory awareness subscale on Months 3, 9, 24, and 36.

It is informative to compare the effect of seizures on children’s developmental trajectories with effects of sleep problems and high-TV use (Table 5). On the combinatorial language subscale, the negative effect of seizures (LS Mean difference=0.43, SE=1.38; P-value=0.753) and sleep problems (1.63, 1.28; 0.2022) on children’s development was relatively smaller that the effect of high-TV use (2.58, 1.04; 0.0128). On the expressive language subscale, seizures (2.61, 0.9; 0.0037) had greater negative effect than sleep problems (1.62, 0.94; 0.0856); while high-TV use had a positive effect (−1.26, 0.7; 0.0719). On the sociability subscale, seizures (2.58, 1.2; 0.0320) had similar negative effect to that of sleep problems (2.67, 1.31; 0.0426) and to high-TV use (1.82, 0.99; 0.0663). On the sensory awareness subscale, seizures (2.97, 1.05; 0.0047) had greater negative effect than that of sleep problems (1.51, 1.11; 0.1751) and high-TV use (1.58, 0.85; 0.0631). On the Health subscale, seizures were associated with significantly poor health outcome (8.29, 2.02; <0.0001) that was comparable with the negative effect of sleep problems (14.14, 1.93; <0.0001); high-TV use had no effect on the health trajectory (1.05,1.52; 0.4898).

**Table 5.**
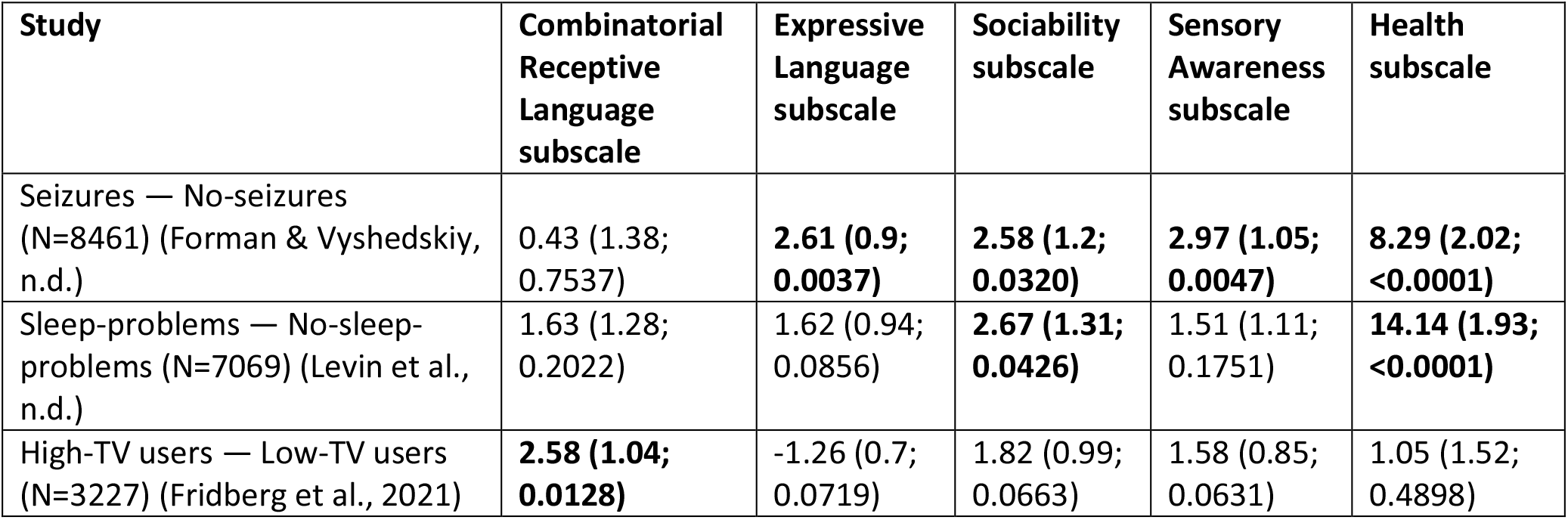
Absolute score difference between groups at the end of three-year study period (Month 36). For every study, children in both groups were matched by age, gender, receptive language, expressive language, sociability, sensory awareness, and health at the 1^st^ evaluation (baseline) using propensity score analysis. Children’s age at baseline in all studies was 2 to 5 years. The differences between groups are presented as: LS Mean (SE; P-value).

| Study | Combinatorial<br>Receptive<br>Language<br>subscale | Expressive<br>Language<br>subscale | Sociability<br>subscale | Sensory<br>Awareness<br>subscale | Health<br>subscale |
| --- | --- | --- | --- | --- | --- |
| Seizures — No-seizures<br>(N=8461) (Forman & Vyshedskiy,<br>n.d.) | 0.43 (1.38;<br>0.7537) | **2.61 (0.9;**<br>**0.0037)** | **2.58 (1.2;**<br>**0.0320)** | **2.97 (1.05;**<br>**0.0047)** | **8.29 (2.02;**<br>**<0.0001)** |
| Sleep-problems — No-sleep-problems (N=7069) (Levin et al.,<br>n.d.) | 1.63 (1.28;<br>0.2022) | 1.62 (0.94;<br>0.0856) | **2.67 (1.31;**<br>**0.0426)** | 1.51 (1.11;<br>0.1751) | **14.14 (1.93;**<br>**<0.0001)** |
| High-TV users — Low-TV users<br>(N=3227) (Fridberg et al., 2021) | **2.58 (1.04;**<br>**0.0128)** | -1.26 (0.7;<br>0.0719) | 1.82 (0.99;<br>0.0663) | 1.58 (0.85;<br>0.0631) | 1.05 (1.52;<br>0.4898) |

These findings add significantly to the results of previous studies due to the longitudinal nature of the collected data, as the vast majority of studies are cross-sectional. Looking at the changes in the score over three years provides an important tool for systematic approach to the development of treatment strategies. Future studies could utilize the longitudinal approach to analyze the impacts of antiepileptic medications as well various developmental interventions.

## Data Availability

All data produced in the present study are available upon reasonable request to the authors

## Acknowledgements

We wish to thank Dr. Petr Ilyinskii for his scrupulous editing of this manuscript.

## Author contributions

AV and EK designed the study. AV and PF analyzed the data. PF and AV wrote the paper.

## Informed Consent

Caregivers have consented to anonymized data analysis and publication of the results. The study was conducted in compliance with the Declaration of Helsinki (Association, 2013).

## Compliance with Ethical Standards

Using the Department of Health and Human Services regulations found at 45 CFR 46.101(b)(4), the Biomedical Research Alliance of New York LLC Institutional Review Board (IRB) determined that this research project is exempt from IRB oversight.

## Data Availability

De-identified raw data from this manuscript are available from the corresponding author upon reasonable request.

## Code availability statement

Code is available from the corresponding author upon reasonable request.

