## Supplementary Material for "Effect of seizures on developmental trajectories in children with autism"

Lauren Cooper ^1^, Edward Khokhlovich ^3^ and Andrey Vyshedskiy ^1,2,^*

**Table S1: LS Means (SE; 95% CI) for Receptive Language MSEC subscale score. The difference between no Seizures Group and Seizures Group (noSG – SG) is presented as: LS Mean (SE; P-value). The negative noSG – SG difference indicates that the SP group had higher score and therefore more severe symptoms.**

| **Visit Number** | **No Seizures Group** | **Seizures Group** | **noSG – SG** |
| --- | --- | --- | --- |
| Baseline | 28 (0.246; 27.5 - 28.5) | 26.5 (0.261; 26 - 27) | **1.52 (0.34; <0.0001)** |
| Month 6 | 28.2 (0.299; 27.6 - 28.8) | 23.8 (0.316; 23.2 - 24.5) | **4.37 (0.42; <0.0001)** |
| Month 9 | 26.4 (0.313; 25.8 - 27) | 24.1 (0.33; 23.5 - 24.7) | **2.27 (0.44; <0.0001)** |
| Month 12 | 26.3 (0.352; 25.6 - 27) | 23.2 (0.37; 22.5 - 23.9) | **3.13 (0.5; <0.0001)** |
| Month 15 | 25.6 (0.405; 24.8 - 26.4) | 22.8 (0.433; 22 - 23.7) | **2.81 (0.58; <0.0001)** |
| Month 18 | 25.4 (0.486; 24.4 - 26.3) | 22 (0.528; 21 - 23.1) | **3.32 (0.71; <0.0001)** |
| Month 21 | 24.4 (0.559; 23.3 - 25.5) | 22.6 (0.566; 21.4 - 23.7) | **1.81 (0.78; 0.021)** |
| Month 24 | 23.2 (0.606; 22 - 24.4) | 23.9 (0.633; 22.7 - 25.1) | -0.74 (0.87; 0.3933) |
| Month 27 | 22.9 (0.672; 21.6 - 24.2) | 22.3 (0.724; 20.9 - 23.8) | 0.52 (0.98; 0.5934) |
| Month 30 | 22.6 (0.754; 21.1 - 24.1) | 21.4 (0.839; 19.8 - 23.1) | 1.19 (1.12; 0.2886) |
| Month 33 | 22.8 (0.917; 21 - 24.6) | 23.2 (0.947; 21.4 - 25.1) | -0.44 (1.31; 0.7347) |
| Month 36 | 21.9 (0.949; 20.1 - 23.8) | 22.4 (1.007; 20.4 - 24.3) | -0.43 (1.38; 0.7537) |
| Month 36 - Baseline | -6.07 (0.96; <0.0001) | -4.12 (1.01; <0.0001) | na |

**Table S2: LS Means (SE; 95% CI) for Expressive Language measured by the Subscale 1 of ATEC. The difference between no Seizures Group and Seizures Group (noSG – SG) is presented as: LS Mean (SE; P-value). The negative noSG – SG difference indicates that the SP group had higher score and therefore more severe symptoms.**

| **Visit Number** | **No Seizures Group** | **Seizures Group** | **noSG – SG** |
| --- | --- | --- | --- |
| Baseline | 16.23 (0.165; 15.91 - 16.6) | 17.8 (0.188; 17.43 - 18.2) | 0.41 (0.23; 0.0725) |
| Month 6 | 14.7 (0.199; 14.31 - 15.1) | 16.1 (0.221; 15.68 - 16.5) | 0.23 (0.28; 0.4062) |
| Month 9 | 13.73 (0.208; 13.32 - 14.1) | 15.5 (0.226; 15.08 - 16) | **-0.62 (0.29; 0.0328)** |
| Month 12 | 13.14 (0.233; 12.68 - 13.6) | 15.2 (0.259; 14.65 - 15.7) | **-0.75 (0.33; 0.0217)** |
| Month 15 | 12.4 (0.267; 11.88 - 12.9) | 14.2 (0.299; 13.6 - 14.8) | **-0.84 (0.38; 0.0275)** |
| Month 18 | 11.58 (0.32; 10.96 - 12.2) | 13.9 (0.355; 13.17 - 14.6) | **-1.98 (0.46; <0.0001)** |
| Month 21 | 11.32 (0.367; 10.6 - 12) | 13.5 (0.399; 12.67 - 14.2) | **-1.68 (0.51; 0.0011)** |
| Month 24 | 10.8 (0.398; 10.02 - 11.6) | 13.1 (0.473; 12.14 - 14) | **-1.6 (0.57; 0.0049)** |
| Month 27 | 10.38 (0.44; 9.52 - 11.2) | 12.6 (0.496; 11.6 - 13.5) | **-2.61 (0.64; <0.0001)** |
| Month 30 | 10.29 (0.494; 9.32 - 11.3) | 12.4 (0.546; 11.33 - 13.5) | -1.41 (0.73; 0.0546) |
| Month 33 | 9.88 (0.599; 8.7 - 11.1) | 11 (0.623; 9.76 - 12.2) | **-3 (0.86; 0.0005)** |
| Month 36 | 9.02 (0.619; 7.8 - 10.2) | 11.8 (0.748; 10.3 - 13.2) | **-2.61 (0.9; 0.0037)** |
| Month 36 - Baseline | -7.21 (0.62; <0.0001) | -4.2 (0.66; <0.0001) | na |

**Table S3: LS Means (SE; 95% CI) for Sociability subscale score measured by the Subscale 2 of ATEC. The difference between no Seizures Group and Seizures Group (noSG – SG) is presented as: LS Mean (SE; P-value). The negative noSG – SG difference indicates that the SP group had higher score and therefore more severe symptoms.**

| **Visit Number** | **No Seizures Group** | **Seizures Group** | **noSG – SG** |
| --- | --- | --- | --- |
| Baseline | 17 (0.207; 16.62 - 17.4) | 16.5 (0.22; 16.08 - 16.9) | 0.52 (0.29; 0.0692) |
| Month 6 | 15.5 (0.254; 15.02 - 16) | 16.5 (0.269; 15.95 - 17) | **-0.95 (0.36; 0.0078)** |
| Month 9 | 14.9 (0.266; 14.36 - 15.4) | 16.6 (0.28; 16.1 - 17.2) | **-1.77 (0.37; <0.0001)** |
| Month 12 | 14.7 (0.301; 14.11 - 15.3) | 16 (0.316; 15.36 - 16.6) | **-1.28 (0.43; 0.0026)** |
| Month 15 | 14.2 (0.348; 13.47 - 14.8) | 16.3 (0.372; 15.55 - 17) | **-2.13 (0.5; <0.0001)** |
| Month 18 | 13.9 (0.42; 13.12 - 14.8) | 16.4 (0.456; 15.48 - 17.3) | **-2.42 (0.61; <0.0001)** |
| Month 21 | 13.4 (0.484; 12.5 - 14.4) | 16.1 (0.49; 15.18 - 17.1) | **-2.7 (0.68; <0.0001)** |
| Month 24 | 14 (0.526; 12.93 - 15) | 17 (0.548; 15.88 - 18) | **-2.99 (0.75; <0.0001)** |
| Month 27 | 13.4 (0.584; 12.27 - 14.6) | 17.1 (0.628; 15.91 - 18.4) | **-3.73 (0.85; <0.0001)** |
| Month 30 | 13.2 (0.655; 11.94 - 14.5) | 16.1 (0.729; 14.67 - 17.5) | **-2.87 (0.97; 0.0032)** |
| Month 33 | 13.4 (0.799; 11.81 - 14.9) | 14.8 (0.825; 13.18 - 16.4) | -1.42 (1.14; 0.2134) |
| Month 36 | 11.5 (0.827; 9.87 - 13.1) | 14.1 (0.877; 12.34 - 15.8) | **-2.58 (1.2; 0.0320)** |
| Month 36 - Baseline | -5.54 (0.84; <0.0001) | -2.44 (0.88; 0.0058) | na |

**Table S4: LS Means (SE; 95% CI) for the Sensory/Cognitive Awareness subscale score measured by the Subscale 3 of ATEC. The difference between no Seizures Group and Seizures Group (noSG – SG) is presented as: LS Mean (SE; P-value). The negative noSG – SG difference indicates that the SP group had higher score and therefore more severe symptoms.**

| **Visit Number** | **No Seizures Group** | **Seizures Group** | **noSG – SG** |
| --- | --- | --- | --- |
| Baseline | 17.1 (0.182; 16.8 - 17.5) | 16.3 (0.194; 15.9 - 16.7) | **0.84 (0.25; 0.0008)** |
| Month 6 | 15.8 (0.223; 15.4 - 16.3) | 16.1 (0.236; 15.6 - 16.6) | -0.27 (0.31; 0.3899) |
| Month 9 | 15.2 (0.233; 14.8 - 15.7) | 16.1 (0.247; 15.6 - 16.6) | **-0.9 (0.33; 0.0061)** |
| Month 12 | 14.8 (0.264; 14.3 - 15.3) | 15.2 (0.277; 14.6 - 15.7) | -0.39 (0.37; 0.2934) |
| Month 15 | 14.7 (0.304; 14.1 - 15.3) | 15.2 (0.326; 14.5 - 15.8) | -0.43 (0.44; 0.3289) |
| Month 18 | 14.2 (0.367; 13.5 - 14.9) | 15.2 (0.399; 14.4 - 16) | -1.05 (0.53; 0.05) |
| Month 21 | 14.1 (0.423; 13.3 - 14.9) | 15 (0.429; 14.1 - 15.8) | -0.88 (0.59; 0.1389) |
| Month 24 | 13.3 (0.46; 12.4 - 14.2) | 15.6 (0.48; 14.6 - 16.5) | **-2.29 (0.66; 0.0005)** |
| Month 27 | 13.1 (0.51; 12.1 - 14.1) | 14.7 (0.549; 13.6 - 15.7) | -1.56 (0.74; 0.0365) |
| Month 30 | 13.6 (0.573; 12.5 - 14.7) | 15.5 (0.638; 14.2 - 16.7) | **-1.87 (0.85; 0.0282)** |
| Month 33 | 12.1 (0.698; 10.7 - 13.4) | 13.5 (0.721; 12.1 - 14.9) | -1.46 (1; 0.1433) |
| Month 36 | 12.6 (0.722; 11.2 - 14) | 15.5 (0.766; 14 - 17) | **-2.97 (1.05; 0.0047)** |
| Month 36 - Baseline | -4.54 (0.73; <0.0001) | -0.73 (0.77; 0.3419) | na |

**Table S5: LS Means (SE; 95% CI) for Health/Physical/Behavior subscale score measured by the Subscale 4 of ATEC. The difference between no Seizures Group and Seizures Group (noSG – SG) is presented as: LS Mean (SE; P-value). The negative noSG – SG difference indicates that the SP group had higher score and therefore more severe symptoms.**

| **Visit Number** | **No Seizures Group** | **Seizures Group** | **noSG – SG** |
| --- | --- | --- | --- |
| Baseline | 28.7 (0.352; 28 - 29.4) | 28.4 (0.375; 27.7 - 29.2) | 0.25 (0.49; 0.6033) |
| Month 6 | 25 (0.432; 24.1 - 25.8) | 29.5 (0.457; 28.7 - 30.4) | **-4.58 (0.61; <0.0001)** |
| Month 9 | 24.3 (0.452; 23.4 - 25.2) | 30.2 (0.477; 29.2 - 31.1) | **-5.87 (0.64; <0.0001)** |
| Month 12 | 25 (0.511; 24 - 26) | 30.2 (0.537; 29.1 - 31.2) | **-5.19 (0.72; <0.0001)** |
| Month 15 | 24.1 (0.589; 22.9 - 25.2) | 29 (0.631; 27.8 - 30.2) | **-4.93 (0.84; <0.0001)** |
| Month 18 | 23.9 (0.709; 22.5 - 25.3) | 30.3 (0.773; 28.8 - 31.9) | **-6.47 (1.03; <0.0001)** |
| Month 21 | 22.4 (0.817; 20.8 - 24) | 30.8 (0.829; 29.2 - 32.4) | **-8.39 (1.15; <0.0001)** |
| Month 24 | 21.8 (0.888; 20 - 23.5) | 29.4 (0.927; 27.6 - 31.2) | **-7.61 (1.27; <0.0001)** |
| Month 27 | 23.1 (0.984; 21.2 - 25.1) | 28.5 (1.061; 26.4 - 30.6) | **-5.38 (1.43; 0.0002)** |
| Month 30 | 22.1 (1.105; 20 - 24.3) | 28.6 (1.231; 26.2 - 31.1) | **-6.5 (1.64; <0.0001)** |
| Month 33 | 22 (1.347; 19.4 - 24.7) | 31.8 (1.392; 29 - 34.5) | **-9.71 (1.92; <0.0001)** |
| Month 36 | 19.7 (1.393; 17 - 22.4) | 28 (1.479; 25.1 - 30.9) | **-8.29 (2.02; <0.0001)** |
| Month 36 - Baseline | -8.99 (1.41; <0.0001) | -0.45 (1.49; 0.7633) | na |

**Table S6: LS Means (SE; 95% CI) for Combinatorial Receptive Language measured by the MSEC subscale when controlling for the health score. The difference between no Seizures Group and Seizures Group (noSG – SG) is presented as: LS Mean (SE; P-value). The negative noSG – SG difference indicates that the SP group had higher score and therefore more severe symptoms.**

| **Visit Number** | **No Seizures Group** | **Seizures Group** | **noSG – SG** |
| --- | --- | --- | --- |
| Baseline | 26.9 (0.286; 26.3 - 27.4) | 25.2 (0.298; 24.7 - 25.8) | **1.62 (0.33; <0.0001)** |
| Month 6 | 26.5 (0.332; 25.9 - 27.2) | 22.8 (0.343; 22.2 - 23.5) | **3.71 (0.41; <0.0001)** |
| Month 9 | 24.6 (0.346; 24 - 25.3) | 23.2 (0.353; 22.5 - 23.9) | **1.44 (0.43; 0.0009)** |
| Month 12 | 24.6 (0.38; 23.8 - 25.3) | 22.3 (0.392; 21.5 - 23.1) | **2.27 (0.49; <0.0001)** |
| Month 15 | 23.9 (0.428; 23 - 24.7) | 21.7 (0.453; 20.8 - 22.6) | **2.19 (0.57; 0.0001)** |
| Month 18 | 23.6 (0.503; 22.7 - 24.6) | 21.1 (0.537; 20.1 - 22.2) | **2.54 (0.69; 0.0002)** |
| Month 21 | 22.3 (0.573; 21.2 - 23.5) | 21.6 (0.571; 20.5 - 22.8) | 0.68 (0.77; 0.3763) |
| Month 24 | 21 (0.616; 19.8 - 22.2) | 22.5 (0.635; 21.3 - 23.8) | -1.48 (0.85; 0.0804) |
| Month 27 | 21 (0.676; 19.6 - 22.3) | 21.1 (0.724; 19.7 - 22.5) | -0.14 (0.96; 0.8801) |
| Month 30 | 20.7 (0.757; 19.2 - 22.2) | 20.2 (0.835; 18.5 - 21.8) | 0.54 (1.09; 0.6244) |
| Month 33 | 20.9 (0.914; 19.1 - 22.7) | 22.4 (0.935; 20.6 - 24.2) | -1.5 (1.29; 0.2444) |
| Month 36 | 19.5 (0.944; 17.7 - 21.4) | 21.3 (0.995; 19.4 - 23.3) | -1.78 (1.35; 0.1864) |
| Month 36 - Baseline | -7.31 (0.94; <0.0001) | -3.91 (0.99; <0.0001) | na |

**Table S7: LS Means (SE; 95% CI) for Expressive Language measured by the Subscale 1 of ATEC when controlling for the health score. The difference between no Seizures Group and Seizures Group (noSG – SG) is presented as: LS Mean (SE; P-value). The negative noSG – SG difference indicates that the SP group had higher score and therefore more severe symptoms.**

| **Visit Number** | **No Seizures Group** | **Seizures Group** | **noSG – SG** |
| --- | --- | --- | --- |
| Baseline | 16.24 (0.195; 15.86 - 16.6) | 15.89 (0.204; 15.49 - 16.3) | 0.35 (0.23; 0.1232) |
| Month 6 | 14.76 (0.226; 14.31 - 15.2) | 14.49 (0.233; 14.04 - 15) | 0.26 (0.28; 0.3504) |
| Month 9 | 13.83 (0.235; 13.37 - 14.3) | 14.43 (0.24; 13.96 - 14.9) | **-0.6 (0.29; 0.042)** |
| Month 12 | 13.25 (0.257; 12.74 - 13.8) | 13.95 (0.265; 13.43 - 14.5) | **-0.71 (0.33; 0.0326)** |
| Month 15 | 12.57 (0.288; 12.01 - 13.1) | 13.34 (0.306; 12.74 - 13.9) | **-0.77 (0.38; 0.0461)** |
| Month 18 | 11.66 (0.338; 11 - 12.3) | 13.65 (0.361; 12.94 - 14.4) | **-1.99 (0.46; <0.0001)** |
| Month 21 | 11.53 (0.384; 10.78 - 12.3) | 13.07 (0.384; 12.32 - 13.8) | **-1.54 (0.52; 0.0029)** |
| Month 24 | 11 (0.413; 10.19 - 11.8) | 12.41 (0.426; 11.58 - 13.2) | **-1.42 (0.57; 0.0129)** |
| Month 27 | 10.44 (0.453; 9.55 - 11.3) | 13.03 (0.485; 12.08 - 14) | **-2.59 (0.64; <0.0001)** |
| Month 30 | 10.41 (0.507; 9.42 - 11.4) | 11.76 (0.559; 10.66 - 12.9) | -1.34 (0.73; 0.0666) |
| Month 33 | 9.96 (0.611; 8.76 - 11.2) | 12.92 (0.625; 11.7 - 14.1) | **-2.97 (0.86; 0.0006)** |
| Month 36 | 9.42 (0.631; 8.18 - 10.7) | 11.73 (0.665; 10.43 - 13) | **-2.32 (0.9; 0.0103)** |
| Month 36 - Baseline | -6.83 (0.63; <0.0001) | -4.16 (0.66; <0.0001) | na |

**Table S8: LS Means (SE; 95% CI) for Sociability measured by the Subscale 2 of ATEC when controlling for the health score. The difference between no Seizures Group and Seizures Group (noSG – SG) is presented as: LS Mean (SE; P-value). The negative noSG – SG difference indicates that the SP group had higher score and therefore more severe symptoms.**

| **Visit Number** | **No Seizures Group** | **Seizures Group** | **noSG – SG** |
| --- | --- | --- | --- |
| Baseline | 18.1 (0.237; 17.6 - 18.5) | 17.7 (0.246; 17.3 - 18.2) | 0.33 (0.28; 0.2359) |
| Month 6 | 17.2 (0.277; 16.6 - 17.7) | 17.5 (0.285; 16.9 - 18.1) | -0.33 (0.34; 0.3435) |
| Month 9 | 16.7 (0.288; 16.1 - 17.3) | 17.5 (0.293; 17 - 18.1) | **-0.84 (0.36; 0.0207)** |
| Month 12 | 16.4 (0.318; 15.8 - 17) | 16.9 (0.327; 16.3 - 17.5) | -0.49 (0.41; 0.2341) |
| Month 15 | 16 (0.359; 15.3 - 16.7) | 17.4 (0.38; 16.6 - 18.1) | **-1.38 (0.48; 0.0039)** |
| Month 18 | 15.9 (0.423; 15.1 - 16.8) | 17.4 (0.452; 16.5 - 18.3) | **-1.48 (0.58; 0.0112)** |
| Month 21 | 15.6 (0.483; 14.6 - 16.5) | 17.1 (0.482; 16.1 - 18) | **-1.52 (0.65; 0.0194)** |
| Month 24 | 16.3 (0.52; 15.2 - 17.3) | 18.3 (0.536; 17.2 - 19.3) | **-1.99 (0.72; 0.0056)** |
| Month 27 | 15.6 (0.572; 14.4 - 16.7) | 18.4 (0.612; 17.2 - 19.6) | **-2.83 (0.81; 0.0005)** |
| Month 30 | 15.5 (0.641; 14.2 - 16.8) | 17.3 (0.706; 15.9 - 18.7) | **-1.82 (0.93; 0.0497)** |
| Month 33 | 15.9 (0.775; 14.4 - 17.4) | 15.9 (0.793; 14.3 - 17.4) | 0.02 (1.09; 0.9867) |
| Month 36 | 14.4 (0.8; 12.8 - 15.9) | 15.6 (0.844; 13.9 - 17.2) | -1.2 (1.14; 0.294) |
| Month 36 - Baseline | -3.71 (0.8; <0.0001) | -2.18 (0.84; 0.0097) | na |

**Table S9: LS Means (SE; 95% CI) for Sensory Awareness measured by the Subscale 3 of ATEC when controlling for the health score. The difference between no Seizures Group and Seizures Group (noSG – SG) is presented as: LS Mean (SE; P-value). The negative noSG – SG difference indicates that the SP group had higher score and therefore more severe symptoms.**

| **Visit Number** | **No Seizures Group** | **Seizures Group** | **noSG – SG** |
| --- | --- | --- | --- |
| Baseline | 16.9 (0.216; 16.5 - 17.4) | 16.1 (0.226; 15.7 - 16.6) | **0.83 (0.25; 0.001)** |
| Month 6 | 15.7 (0.253; 15.2 - 16.2) | 16 (0.261; 15.5 - 16.5) | -0.24 (0.31; 0.4369) |
| Month 9 | 15.2 (0.263; 14.6 - 15.7) | 16 (0.269; 15.5 - 16.5) | **-0.85 (0.33; 0.0097)** |
| Month 12 | 14.7 (0.29; 14.1 - 15.3) | 15 (0.3; 14.4 - 15.6) | -0.32 (0.37; 0.3956) |
| Month 15 | 14.8 (0.328; 14.2 - 15.4) | 15.1 (0.348; 14.4 - 15.7) | -0.26 (0.44; 0.549) |
| Month 18 | 14.3 (0.387; 13.5 - 15) | 15.2 (0.414; 14.4 - 16) | -0.96 (0.53; 0.0708) |
| Month 21 | 14.2 (0.442; 13.3 - 15) | 14.9 (0.441; 14 - 15.7) | -0.71 (0.59; 0.2322) |
| Month 24 | 13.3 (0.477; 12.4 - 14.2) | 15.5 (0.491; 14.5 - 16.4) | **-2.16 (0.66; 0.001)** |
| Month 27 | 13.2 (0.524; 12.2 - 14.2) | 14.5 (0.56; 13.4 - 15.6) | -1.28 (0.74; 0.0842) |
| Month 30 | 13.7 (0.587; 12.5 - 14.9) | 15.2 (0.647; 14 - 16.5) | -1.54 (0.85; 0.0688) |
| Month 33 | 12.3 (0.71; 10.9 - 13.6) | 13.7 (0.726; 12.2 - 15.1) | -1.4 (1; 0.1606) |
| Month 36 | 12.5 (0.733; 11.1 - 14) | 15.7 (0.773; 14.2 - 17.2) | **-3.13 (1.05; 0.0029)** |
| Month 36 - Baseline | -4.4 (0.73; <0.0001) | -0.44 (0.77; 0.5652) | na |
